# Investigating the cause of a 2021 winter wave of COVID-19 in a border region in Eastern Germany: a mixed-methods study, August to November 2021

**DOI:** 10.1101/2022.10.15.22281123

**Authors:** Buqing Yi, Alexa Laubner, Marlena Stadtmüller, Eva Patrasová, Lenka Šimůnková, Fabian Rost, Sylke Winkler, Susanne Reinhardt, Andreas Dahl, Alexander H. Dalpke

## Abstract

In winter 2020 and 2021 many countries worldwide experienced a COVID-19 pandemic wave which led to severe burdens on healthcare systems and huge economic losses. Yet, it remains unclear how the winter waves started and many debates are ongoing about actions necessary to prevent future winter waves. In this study we deciphered the dynamic course of a winter wave in 2021 in Saxony, a state in Eastern Germany neighboring Czech Republic and Poland. The information we achieved might help future pandemic prevention.

The dynamic course of the 2021 winter wave in Saxony was investigated through integration of multiple virus genomic epidemiology approaches and functional evaluations of locally circulating variants. Through international collaborations, we performed genomic epidemiology analysis on a weekly base with samples from Saxony and also from one neighbor region in the Czech Republic. Phylogeny analyses were used to track transmission chains, monitor virus genetic changes and identify emerging variants. Phylodynamic approaches have been applied to track the dynamic changes of transmission clusters. For identified local variants of interest, active viruses were isolated and functional evaluations were performed.

Genomic epidemiology studies revealed multiple long-lasting community transmission clusters acting as the major driving forces for the winter wave 2021. Analysis of the dynamic courses of two representative long-lasting community transmission clusters indicated similar dynamic changes. In the first 6-8 weeks, the relevant variant was mainly circulating in a small region among young and middle-aged people; after eight weeks, the ratio of people aged above 60 years in the infected population markedly increased, and the virus got more widely spread to distant regions. On the other hand, the transmission cluster caused by a locally occurring variant showed a different transmission pattern. It got geographically widely distributed within six weeks, with many people aged above 60 years being infected since the beginning of the cluster, indicating a higher risk for escalating healthcare burdens. This variant displayed a relative growth advantage compared to co-circulating Delta sub-lineages. Functional analyses revealed a replication advantage, but no advantage in immune evasion ability.

This study indicated that long-lasting community transmission clusters starting between August and October caused by imported variants as well as locally occurring variants all contributed to the development of the 2021 winter wave in Saxony. In particular, the cluster derived from a locally occurring variant with certain growth advantage might have stressed local healthcare systems.

## Introduction

To understand the impact of the COVID-19 pandemic and to design effective mitigation or prevention strategies, it is critical to decipher the transmissibility, prevalence and patterns of movement of SARS-CoV-2 infections, for which phylogenies have provided key information about the international spread of SARS-CoV-2 and enabled investigation of individual outbreaks and transmission chains ^1,2^. Phylodynamic approaches integrate evolutionary, demographic and epidemiological concepts and play an important role in tracking virus genetic changes, identifying emerging variants and informing public health strategy ^3^. With these two powerful tools, genomic epidemiology may provide a lot of valuable information from several different aspects, from public health to important clinical parameters. To monitor the evolution of SARS-CoV-2 variants and to investigate transmission chains, since 2020 we have performed virus surveillance and genomic epidemiology research in Saxony, one state in Eastern Germany which neighbors the Czech Republic and Poland. Also, through international collaboration, between 2021 March and 2022 March we performed genomic epidemiology analysis on a weekly basis with SARS-CoV-2 samples collected from a border region between Saxony, Poland and the Czech Republic in a global background. For identified virus mutant variants, active viruses were isolated and functional evaluations were performed to test their replication fitness and neutralization sensitivity against vaccine elicited serum neutralizing antibodies. Thereby we previously identified a B.1.1.7 sub-lineage predominant in several European countries, such as Czech Republic, Austria and Slovakia ^4^. In addition to monitor virus evolution and transmission, with genomic epidemiology we also analyzed why and how the pandemic developed in Saxony to evaluate the local pandemic or post-pandemic conditions, which may help to predict what will take place in future.

In summer and early autumn 2021, the 7-day incidence rates of COVID-19 in Saxony were below 10 during most time, which was much lower compared to the average incidence rates in Germany at that time (Figure 1). However, from October on, the incidence rate in Saxony increased dramatically. At the beginning of November, it already exceeded 500, which was two times higher than the average value in Germany at that time, and the severe burdens on the healthcare system resulted in a lockdown. What happened in Saxony in late autumn, i.e. in September and October, that led to the high incidence in winter? We investigated the sources behind the high COVID-19 incidence in Saxony in autumn and winter 2021.

**Figure 1:**
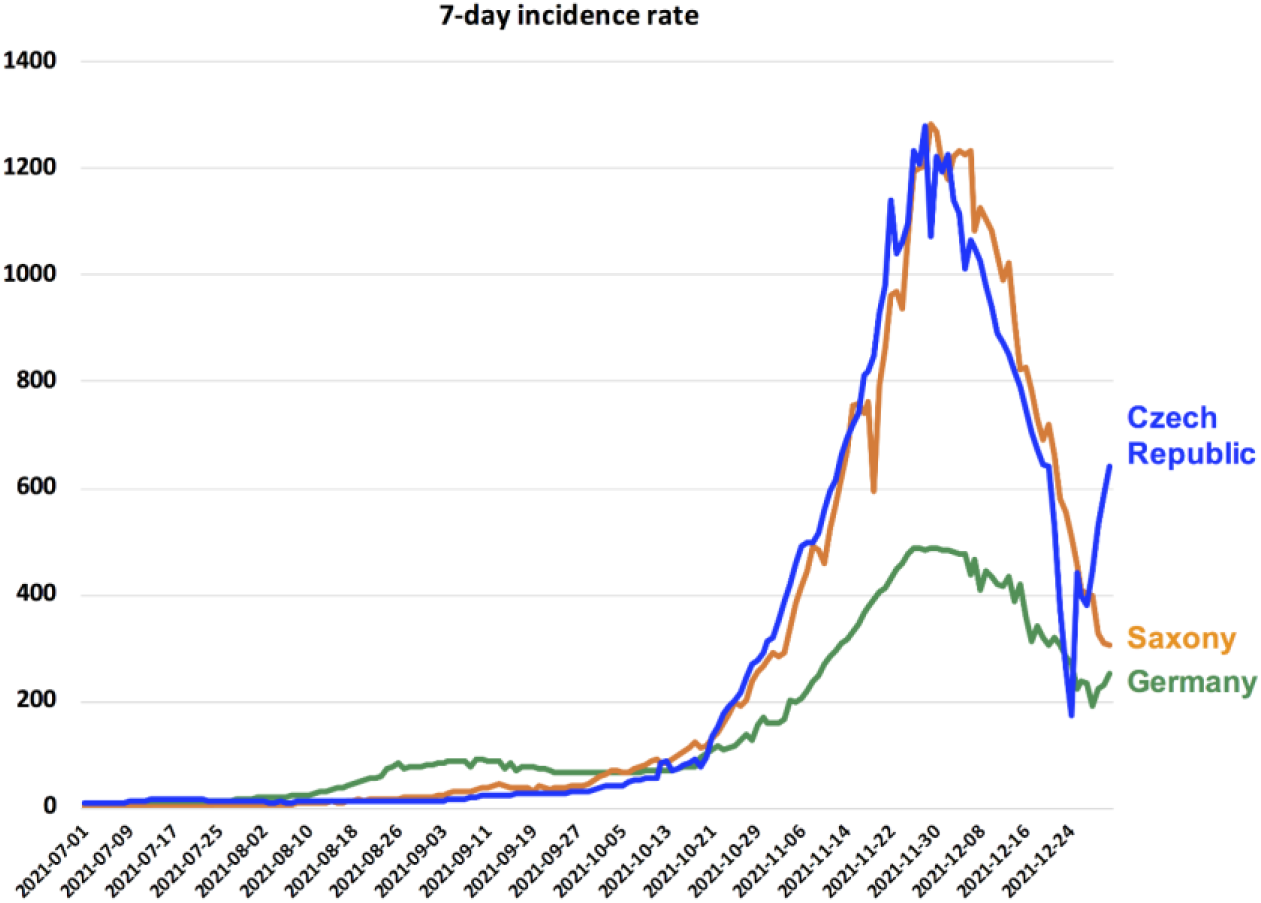
The COVID-19 pandemic waves in late 2021 in Saxony, Germany and the Czech Republic. 7-day incidence rate per 100,000 inhabitants is shown for each region between July and December 2021.

## Methods

### 1. Establishment of genome sequence data set for genomic epidemiology investigation and monitoring emerging variants

We combined SARS-CoV-2 sequences generated from samples collected in a border region between Germany, Poland and Czech Republic, with full-length SARS-CoV-2 sequences periodically downloaded from GISAID ^5^ to build up genome sequence data set for epidemiology investigation and monitoring emerging variants (locally generated sequences were shared on GISAID as well). To detect local transmission clusters, we also performed genomic epidemiology analysis only with self-collected samples with documented meta data. In the German side, usually 5-10% of local SARS-CoV-2 positive samples were sequenced, ranging from 50 to 300 samples per week; In the Czech Republic, usually at least 50 samples per week were sequenced. Geographic coverage of sample collection in this study is shown in supplementary Figure S1. An unbiased sampling procedure was applied to sequencing sample collection without pre-selection of the samples. We performed quality check and filtered out low-quality sequences that met any of the following criteria: 1) sequences with less than 90% genome coverage; 2) genomes with too many private mutations (defined as having >24 mutations relative to the closest sequence in the reference tree); 3) genomes with more than ten ambiguous bases; and 4) genomes with mutation clusters, defined as 6 or more private differences within a 100-nucleotide window. These are the standard quality assessment parameters utilized in NextClade (https://clades.nextstrain.org) ^6^.

### 2. Lineage classification

We used the dynamic lineage classification method through the Phylogenetic Assignment of Named Global Outbreak Lineages (PANGOLIN) software suite (https://github.com/hCoV-2019/pangolin) ^7^. This is intended for identifying the most epidemiologically important lineages of SARS-CoV-2 at the time of analysis ^8^.

### 3. Phylogeny and phylogeographical analyses of SARS-CoV-2

We carried out phylogenetic and phylogeographical analysis to identify transmission clusters, monitor virus genetic changes and infer the transmission routes of AY.36.1 in Europe ^9^ with a custom build of the SARS-CoV-2 NextStrain build (https://github.com/nextstrain/ncov) ^10^. The pipeline includes several Python scripts that manage the analysis workflow. Briefly, it allows for the filtering of genomes, the alignment of genomes in NextClade (https://clades.nextstrain.org) ^6^, phylogenetic tree inference in IQ-Tree ^11^, tree dating ^12^ and ancestral state construction and annotation. For transmission cluster identification, the phylogeny analysis is rooted with “least-squares” methods^13^ to make the phylogeny less affected by square errors of the branch lengths and therefore possibly more accurately reflect sample-to-sample relationship. To infer the transmission routes of AY.36.1 in Europe, only samples fulfilling these criteria on GISAID were included in the analysis: 1. With complete sample collection dates; 2. With a complete sequence (>29,000nt) and less than 5% Ns ; 3. With all the definition mutations of AY.36.1 including signature mutations of AY.36 (S: 1104L, orf1b:721R, orf1b:1538L) and four other AA substitutions: orf9b: 3S, orf3a: 223I, orf1a: 944L, N: 6L. The phylogeny analysis is rooted by *Wuhan*-Hu-1/2019 (GISAID Accession ID: EPI_ISL_402125).

### 4. Phylodynamic analysis of transmission chains

Phylodynamic approaches to track the dynamic changes of transmission clusters were carried out by integrating phylogenetic and demographic information, primarily using RStudio v1.3.1093 with multiple R software, e.g. tidyverse, ggplot and ggmap.

### 5. Epidemiology data

We analyzed daily cases of SARS-CoV-2 in the Czech Republic from publicly released data provided by the Ministry of Health of the Czech Republic (https://onemocneni-aktualne.mzcr.cz/covid-19), and 7-day incidence rates per 100,000 inhabitants were calculated accordingly based on the local population. The data of 7-day-incidence rates per 100,000 inhabitants in Germany or in Saxony were obtained from the Robert Koch Institute (https://www.rki.de/DE/Content/InfAZ/N/Neuartiges_Coronavirus/Fallzahlen.html).

### 6. Relative growth advantage

We analysed SARS-CoV-2 sequences from Germany that were uploaded to GISAID with complete sample collection dates from October 1^st^ to November 30^th^ 2021. A logistic regression model was used to estimate the relative growth advantage of certain variant compared to co-circulating variants as previously reported^14-17^. The model assumes that the increase or decrease of the proportion of a variant follows a logistic function, which is fit to the data by optimizing the maximum likelihood to obtain the logistic growth rate in units per day. Based on that, an estimate of the growth advantage per generation is obtained (assuming the growth advantage arising from a combination of intrinsic transmission advantage, immune evasion, and a prolonged infectious period ^18^, and the relative growth advantage per week (in percentage; 0% means equal growth) is reported. The relative growth advantage estimate reflects the advantage compared to co-circulating variants in the selected country and time frame.

### 7. Viruses

All viruses used were patient isolates cultured from nasopharyngeal swabs. Virus stocks were grown on Vero E6 cells in DMEM GlutaMAX supplemented with 10% FBS, 1% non-essential amino acids and 1% penicillin/streptomycin. The second passage of each virus isolate was used for experiments. The virus isolates BA.1 (hCoV-19/Germany/SN-RKI-I-405124/2021, EPI_ISL_8237557), B.1.1.7 (hCoV-19/Germany/SN-RKI-I-178035/2021, EPI_ISL_2634728), B.1.177 (hCoV-19/Germany/SN-RKI-I-017381/2021, EPI_ISL_1147543), AY.122 (hCoV-19/Germany/SN-RKI-I-348308/2021, EPI_ISL_7101815), AY.36.1 (hCoV-19/Germany/SN-RKI-I-290321/2021, EPI_ISL_5402822), were used in the virus neutralization assay and growth kinetics measurement.

### 8. Virus Neutralization Assay

All sera were derived from healthy individuals following vaccination with triple doses of BNT162b2 (around one month after the third shot). A 2-fold dilution series of each serum was prepared in PBS+ (supplemented with 0.3 % bovine albumin, 1 mM MgCl_2_ and 1 mM CaCl_2_) and each serum concentration was incubated with 50 PFU of AY.36.1, AY.122, BA.1, B.1.1.7 or B.1.177 for 1 h at 37°C. Confluent Vero E6 cells seeded the day before were infected with the virus-containing serum dilutions for 1 h at 37°C and 5% CO_2_ with occasional shaking. The inoculum was aspirated, cells washed with PBS and subsequently overlayed with semi-viscous Avicel Overlay Medium (double-strength DMEM, Avicel RC-581 in H2O 0.75 %, 10 % FCS, 0.01 % DEAE-Dextran and 0.05 % NaHCO_3_). After 3 days, cells were stained with 0.1 % crystal violet in 10 % formaldehyde and plaques were counted. Eleven dilutions of each serum were tested. The neutralization assay was performed in three independent experiments with each serum. Each experiment was conducted in technical duplicates of each serum. Technical duplicates were averaged before further calculations. ID_50_ values were calculated using 4^th^ order nonlinear regression curve fits with GraphPad Prism 9.

### 9. Virus Growth Kinetics

Calu 3 cells were seeded 3 days prior to infection. On the day of infection, cells were infected with AY.122, AY.36.1 or BA.1 at MOI 0.1 diluted in PBS+ (PBS supplemented with 0.3 % BSA and 1 mM MgCl_2_ and 1 mM CaCl_2_) for 1h at 37°C and 5% CO_2_ with occasional shaking. Afterwards, the inoculum was aspirated, the cells were washed with PBS and fresh medium (DMEM GlutaMAX supplemented with 10% FBS, 1 % non-essential amino acids, 1% sodium pyruvate and 1% penicillin/streptomycin) was added. Supernatants were removed at 8, 16, 24, 48 hours post infection (hpi). Infectious virus particles in the supernatant were determined using plaque assay, which was performed analogously to the neutralization assay from the infection step onwards. Results are given as plaque forming units (PFU) per ml. Graphs were generated using GraphPad Prism 9.

## Results

### 1. The spreading of a few specific delta variants accompanied the high incidence rates in a cross-border region between Eastern Germany and the Czech Republic

We first investigated the predominant SARS-CoV-2 variants responsible for the high COVID-19 incidence in Saxony in last autumn and winter. Between October to November 2021, four SARS-CoV-2 Delta sub-lineages were most frequently detected in Saxony: AY.122, AY.43, AY.4 and AY.36, indicating these variants caused most infections during that period (Figure 2A). In the neighbor country Czech Republic, the predominant variants were slightly different, with AY.122, AY.43, AY.4 and AY.4.13 accounting for most infections (the analyses for both places were based on information from GISAID acquired on January 31, 2022).

**Figure 2:**
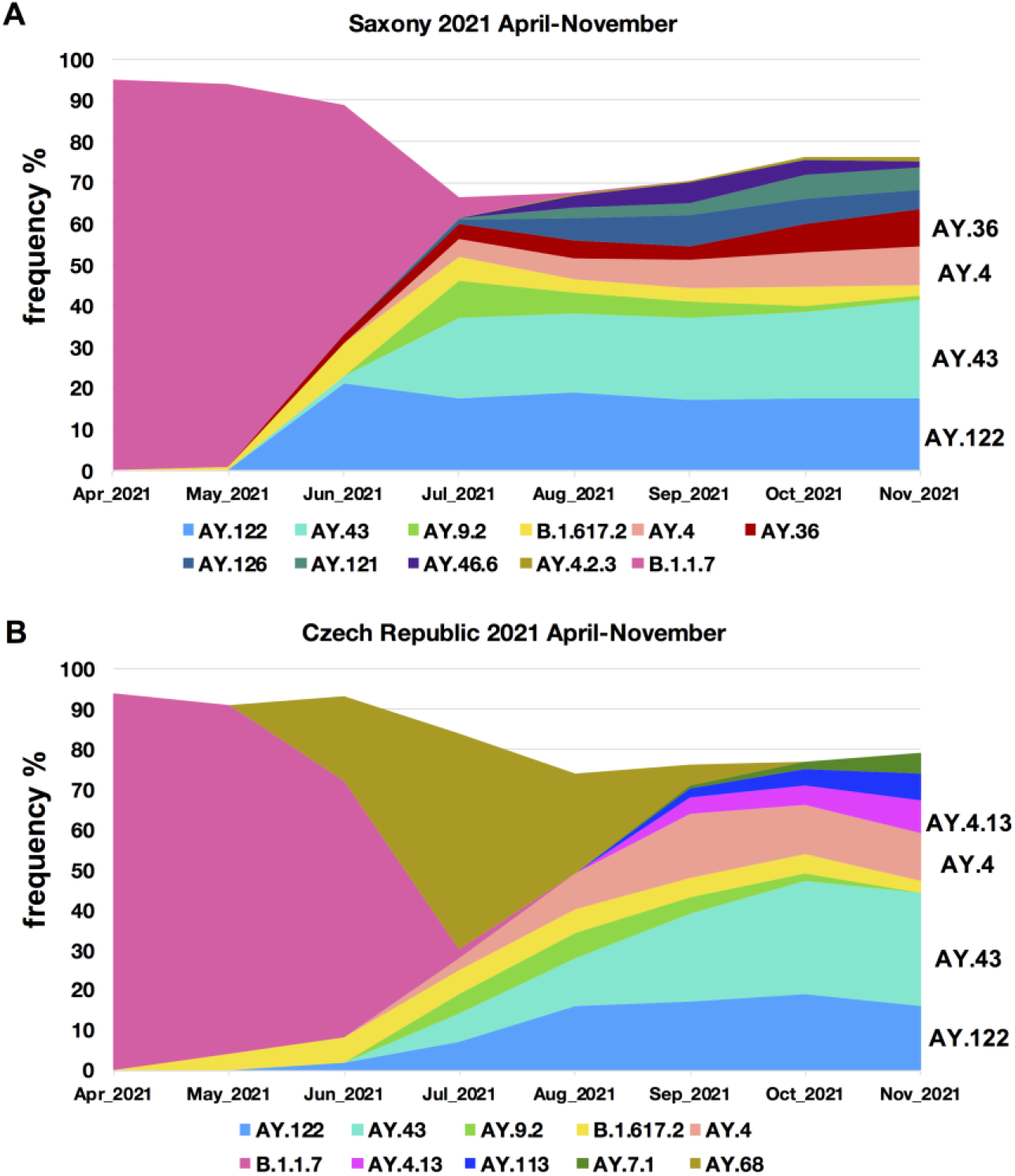
Lineage dynamic changes in Saxony and Czech Republic, April-November 2021. Frequency of detection (%) of each SARS-CoV-2 lineage in each month in **(A)** Saxony and **(B)** Czech Republic is displayed. To achieve a better resolution, lineages with a highest frequency < 5% during this period in each place are not separately shown, while collectively shown as the white blank space in the area plots.

Among these lineages, AY.122 had been reported to be the dominant Delta sub-lineage in Russia and a few other Eastern European countries since April 2021 ^19^. The AY.43 lineage had been detected in multiple European countries since early 2021. AY.4 was distributed worldwide, and also frequently detected in Europe, especially in UK, since April 2021^20^. In Saxony, these several lineages were not frequently detected until late June 2021. This suggested that most cases related with these several lineages (AY.122, AY.43 and AY.4) were caused by either directly imported variants or community transmission clusters derived from the imported variants (if the further transmission of the imported cases had not been prevented).

### 2. Several community transmission clusters of the Delta variant could be identified through genomic epidemiology

With phylogenetic and phylogeographic analyses, genomic epidemiology may serve as a powerful tool for identifying transmission relationships under the condition of lack of contact information ^3^. In this study, through genomic epidemiology analyses, several community transmission clusters were identified in Saxony, including clusters caused by AY.122, AY.43, AY.36 (later named as AY.36.1, with details described in section 4) and a few others, most of which started from August or September 2021 during or shortly after the travel season (mainly the summer holiday season) (sup. Figure 2). Because self-collected samples contained corresponding meta data, we performed further analyses to inspect the details of identified transmission clusters with samples collected till middle of November before the lockdown (Figure 3A). As a representative cluster, the AY.122 cluster kept on from August 2021 through autumn and winter 2021 (Figure 3B&C). From the neighbor region Ustecky Kraj in the Czech Republic, one AY.122 community transmission cluster was also detected, which similarly started from August 2021 (Figure 3D&E).

**Figure 3:**
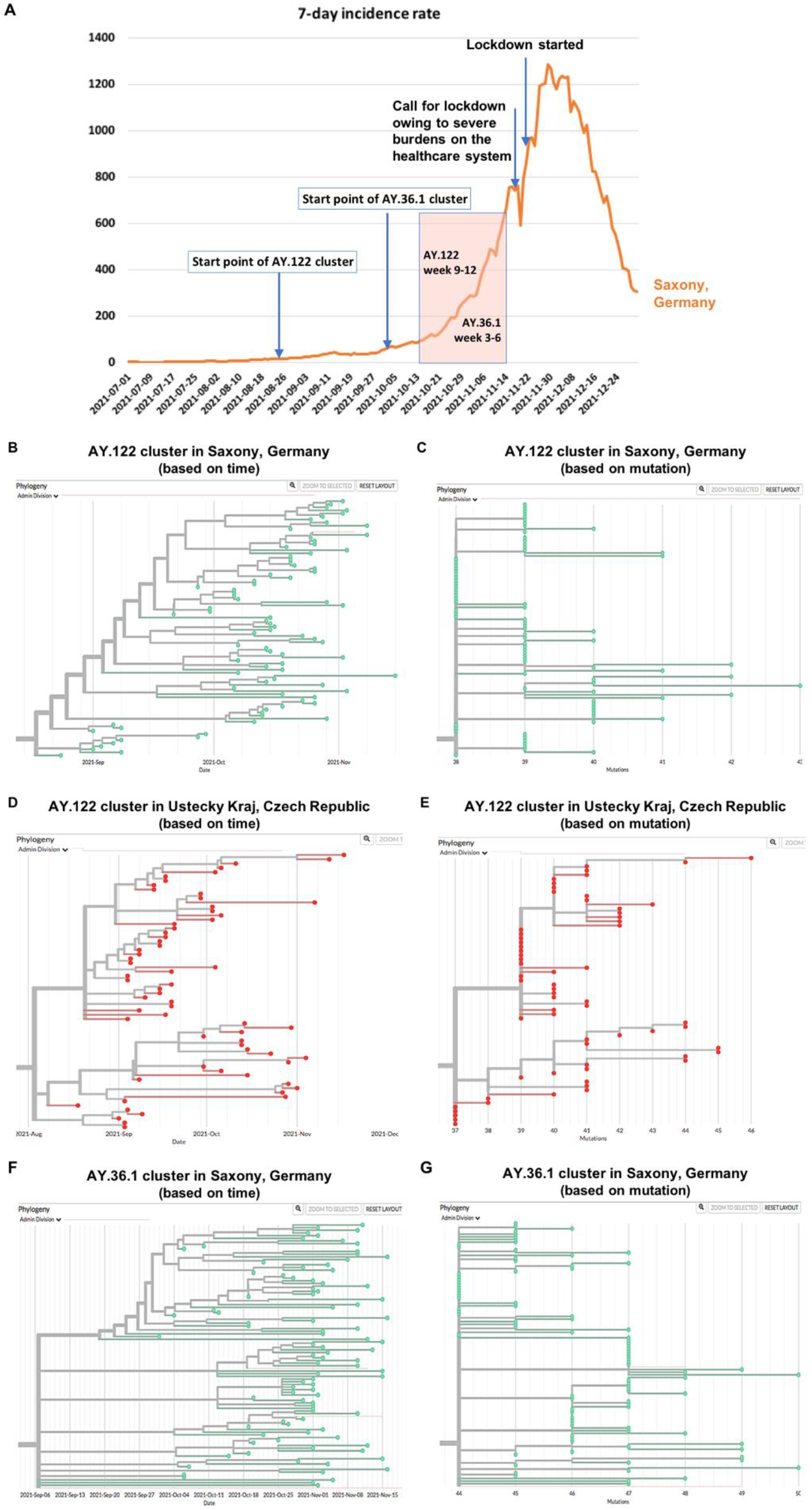
Multiple community transmission clusters were identified through genomic epidemiology, the development course of which was associated with the development course of the winter wave. **(A)** The association of the course of the community transmission clusters with the development course of the winter wave in 2021 in Saxony, Germany. **(B&C)** AY.122 cluster in Saxony, Germany is displayed based on time (B) and mutation (C), respectively. **(D&E)** AY.122 cluster in Ustecky Kraj, Czech Republic is displayed based on time (D) and mutation (E), respectively. **(F&G)** AY.36.1 cluster in Saxony is displayed based on time (F) and mutation (G), respectively. These clusters are displayed with samples collected till middle of November 2021 with each dot representing one sample.

### 3. Dynamic changes of the two AY.122 clusters display a highly similar pattern

With available meta data, we performed more detailed analysis of the AY.122 clusters in both Saxony, Germany, and in Ustecky Kraj, Czech Republic. The samples in one community transmission cluster are often geographically clustered at the early stages of the cluster development^20,21^. Consistent with that, we observed clear geographic clustering in both two AY.122 clusters. As shown in Figure 4A&B, both AY.122 clusters in the two places displayed a similar transmission pattern in the dynamic changes of geographic distribution. In week 1-4, the virus mainly spread locally in a small region; in week 5-8, more people were infected, but still mainly in a small local region; in week 9-12, the virus got more widely spread to the surrounding regions, or even reached further to more remote places such as for the AY.122 cluster in Saxony.

**Figure 4:**
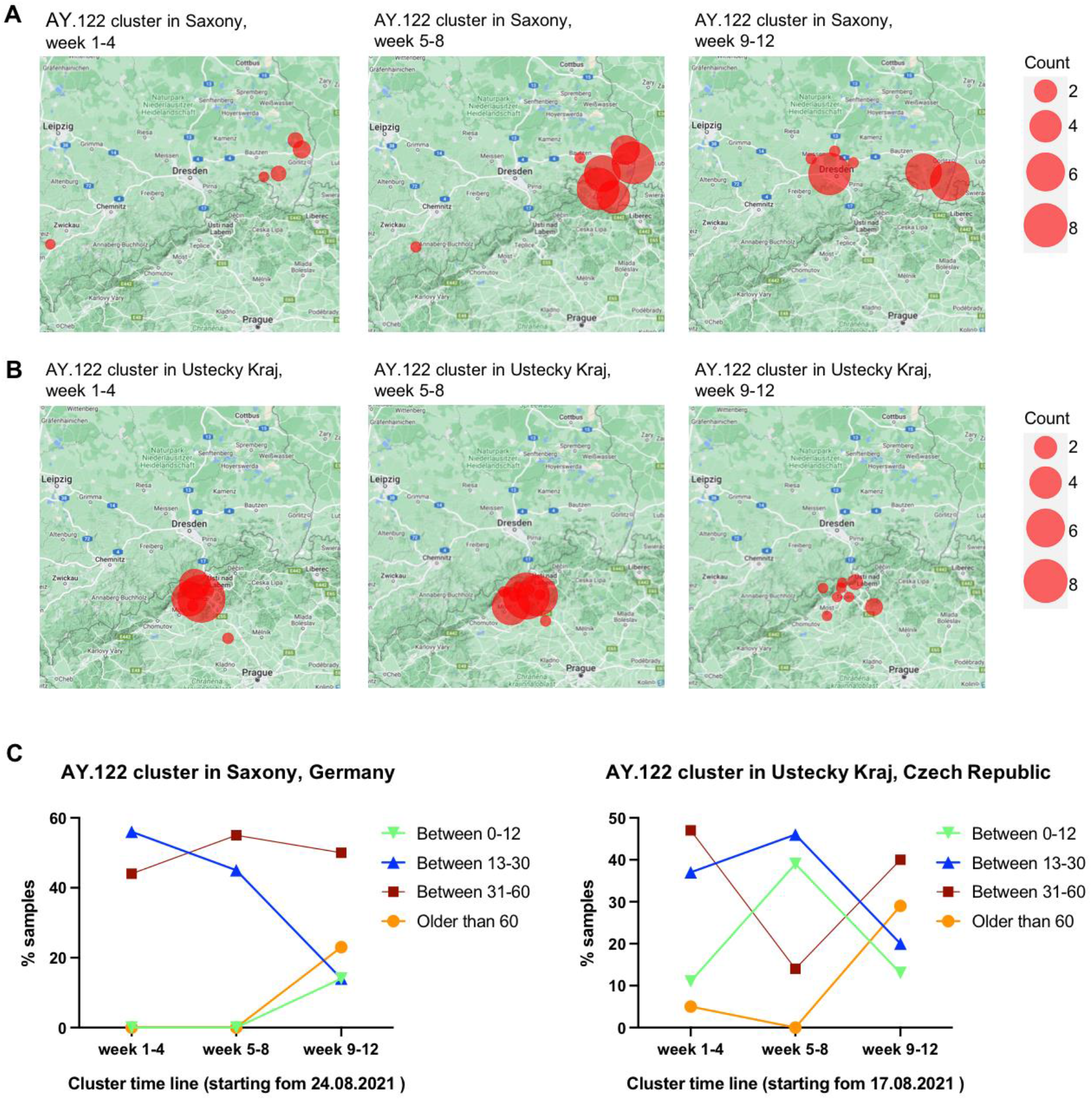
Dynamic changes of geographic distribution and age distribution of detected new samples in each cluster in every four weeks along with the cluster timeline. (A&B). The geographic distribution of detected new samples in the AY.122 cluster in Saxony, Germany (A) and in the AY.122 cluster in Ustecky Kraj, Czech Republic (B) in every four weeks along with the cluster timeline is displayed based on documented postcodes. **(C)**. The age distribution of detected new samples in each cluster in every four weeks is displayed. In both clusters, the ratio of patients aged above 60 years highly increased after 8-week community transmission.

With meta data of patient age, we also analyzed the dynamic changes of age distribution in these two AY.122 clusters. These two clusters also displayed a highly similar pattern in the dynamic change of age destitution (Figure 4C). In week 1-4, mainly young to middle age people (< 60 years old) got infected; in week 5-8, a similar age distribution as week 1-4 was detected; in week 9-12, the ratio of people aged above 60 years clearly increased and went up to about 30% of the newly infected population, which was much higher than that in the first eight weeks (0-5%). This indicated that in both clusters, the ratio of elder patients highly increased after around two-month community transmissions, which means a higher risk for increased healthcare burdens.

The period of week 9-12 of the AY.122 cluster was correlated to the period of the exponential incidence increases between middle of October to the middle of November 2021 in Saxony (as shown in Figure 3A). This observation suggests that the limited spreading of the virus mainly among young to middle age people in the first eight weeks paved a base for the wide spreading of the virus later.

### 4. Community transmissions caused by a locally occurring variant show a distinct transmission pattern

In Saxony, one of the predominant variants responsible for many infection cases was AY.36. Through detailed phylogenetic analyses we discovered that most local AY.36 samples belong to a specific, locally occurring variant derived from virus evolution of AY.36. This special AY.36 grew rapidly in Saxony since October and formed a big cluster. Compared with originally defined AY.36 (with signature mutations S: 1104L, orf1b:721R, orf1b:1538L), most local samples carried four additional AA substitutions: orf9b: 3S, orf3a: 223I, orf1a: 944L, N: 6L. These particular AY.36 samples were later defined as a new Delta sub-lineage AY.36.1 (more detailed information about defining this new lineage can be found at https://github.com/cov-lineages/pango-designation/issues/434). The machine learning process of the PANGOLIN designation included some samples that do not contain all the definition mutations of AY.36.1. In this study, we only focus on AY.36.1 samples that strictly match the definition of AY.36.1 containing both the three AY.36 signature mutations and all the four extra mutations as described above. In the international background, the first AY.36.1 sample was detected in Saxony at the beginning of October, and the earliest 30 samples were almost exclusively from Saxony only with a few exceptions (Figure 5). By end of November, it was already detected in 10 European countries and in several other continents. In December it further spread to multiple other European countries and also to other continents. Regarding transmission route, Germany and Denmark (mainly from December on) were the major source locations of AY.36.1(Figure 5).

**Figure 5:**
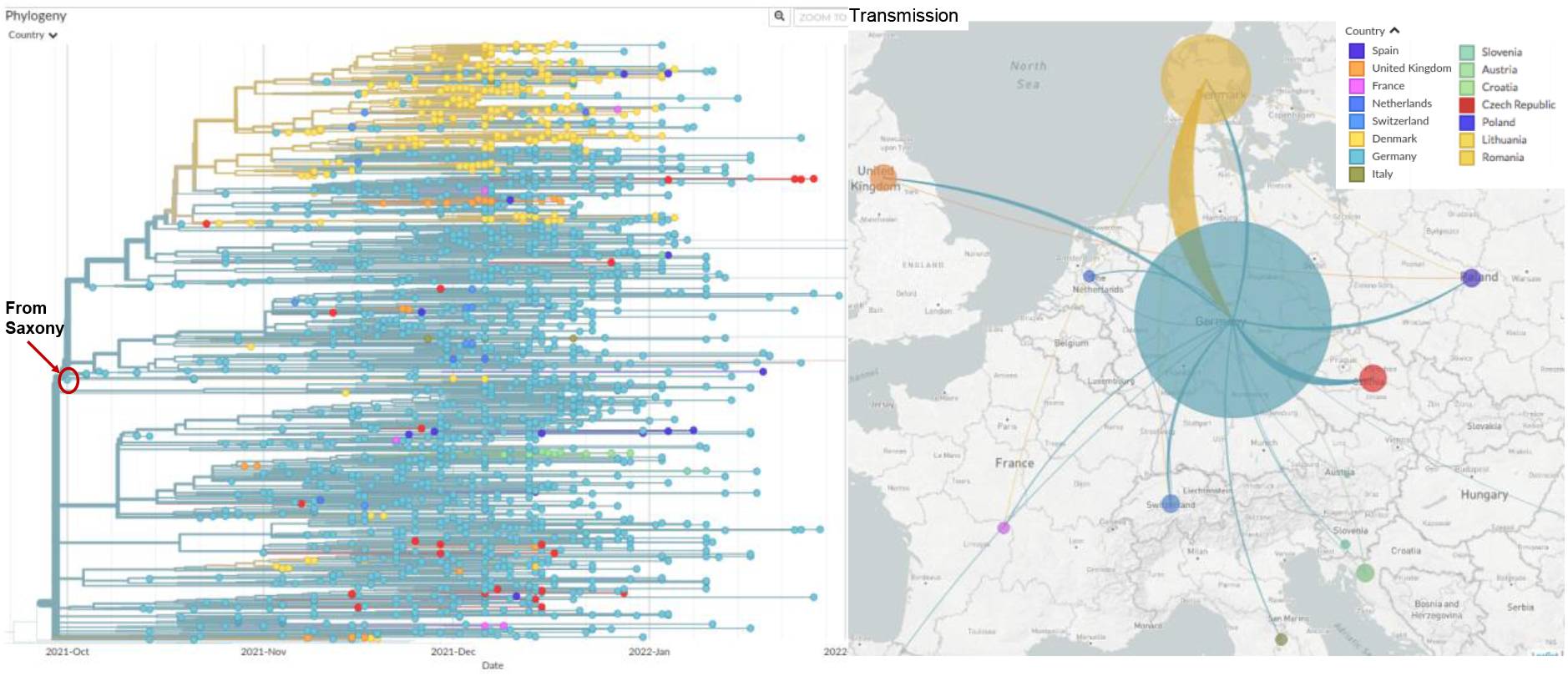
Transmission routes of AY.36.1 in Europe is inferred based on phylogeny analysis. The size of the circle represents the number of genomes from all AY.36.1 in each country collected by end of January 2022. The line colours correspond to the exporting locations. Left: Phylogeny tree of AY.36.1, with branch length representing time. The first sample was detected in Saxony, Germany in early October, 2021; right: estimated transmission routes of AY.36.1 in Europe.

In Saxony, the AY.36.1 variant expanded rapidly and formed a big cluster as shown in Figure 3 F&G. We therefore analyzed whether this locally occurring variant share the same community transmission pattern compared to other Delta variants.

As the AY.36.1 cluster expanded much quicker than the AY.122 cluster, we analyzed the dynamic changes of the transmission pattern of this cluster in every two weeks (instead of every four weeks like that for the AY.122 cluster) starting from the first sample identified at the beginning of October. The period of week 3-6 of the AY.36.1 cluster was correlated to the period of the exponential incidence increases between middle of October to the middle of November 2021 in Saxony (Figure 3A). In the first two weeks, similar to the AY.122 clusters, the AY.36.1 cluster was also mainly spreading in a relatively small local region showing the feature of geographic clustering. But from the 3^rd^ week on, it quickly expanded to the surrounding regions in one explosive way. By end of the 6^th^ week, it was already detected in lots of regions in Saxony (Figure 6A). Regarding age distribution changes, there were many people aged above 60 years being infected by this variant in week 1-2 already, indicating a risk of escalating health care burdens (Figure 6B). These results revealed that the transmission pattern of this cluster caused by the local variant AY.36.1 was different from the transmission pattern of the two AY.122 clusters caused by imported variants.

**Figure 6:**
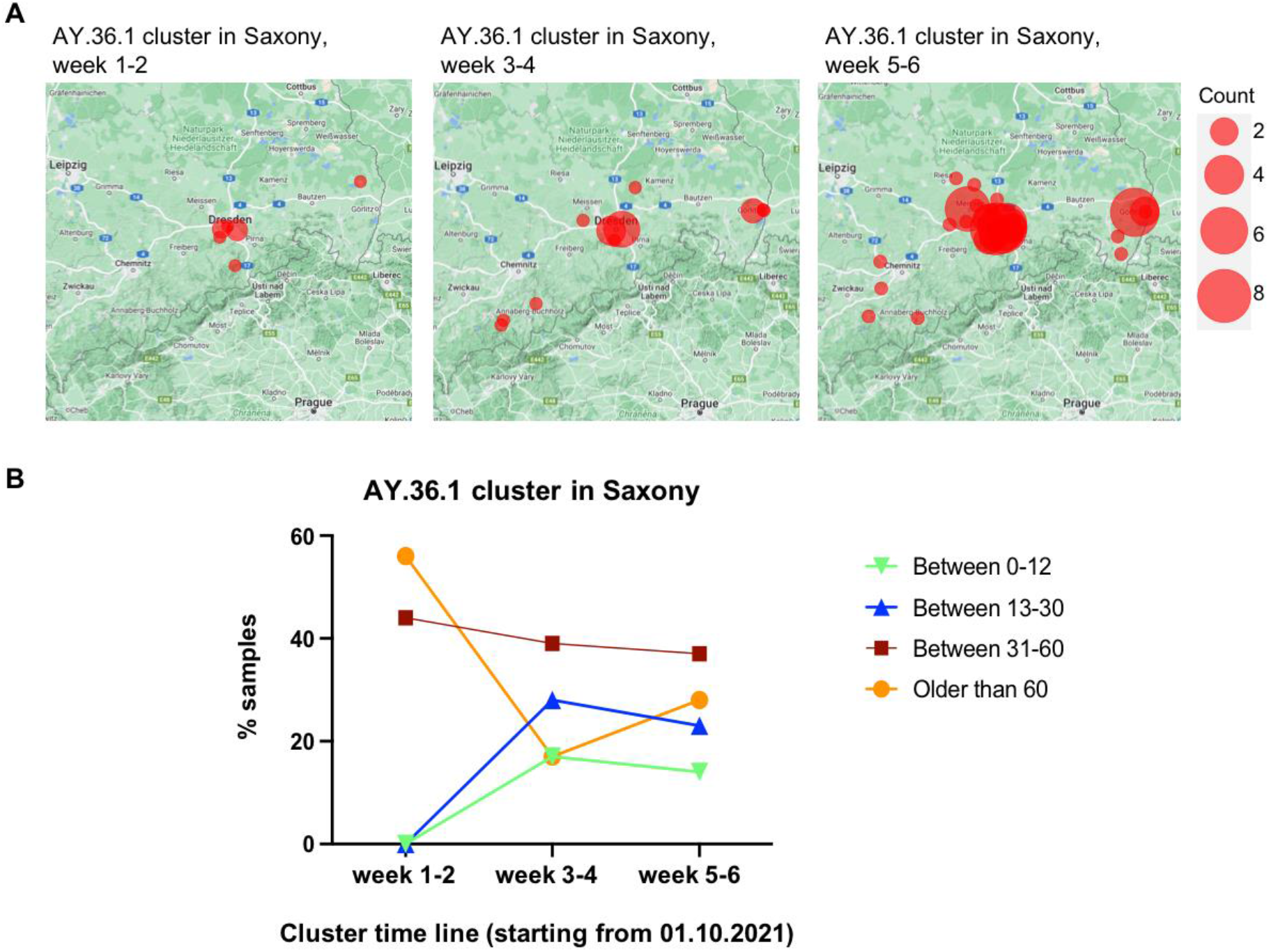
Dynamic changes of geographic distribution and age distribution of detected new samples in the AY.36.1 cluster in every two weeks along with the cluster timeline. **(A)**. The geographic distribution of detected new samples in the AY.36.1 cluster in Saxony, Germany in every two weeks along with the cluster timeline is displayed based on documented postcodes. **(B)**. The age distribution of detected new samples in the AY.36.1 cluster in every two weeks is displayed.

### 5. Comparative analyses of virus propagation and antibody neutralization between AY.36.1 and other VOCs

The estimated growth rate of AY.36.1 in Germany between October and November 2021 was above most other co-circulating Delta sub-lineages, as manifested by a relative growth advantage of the AY.36.1 (sup. Figure 3). To verify if the advantage in growth rate was related to a functional feature of this variant, we evaluated antibody neutralization and virus propagation abilities of this variant in comparison with a few other variants circulating in Germany in 2021. We compared AY.36.1 with another Delta sub-lineage AY.122, the Alpha lineage B.1.1.7, the Omicron sub-lineage BA.1, and one non-VOCs B.1.177, which was one of the predominant lineages during the second wave in winter 2020 and early 2021 ^22^, by testing their susceptibilities to vaccine-elicited serum neutralizing antibodies in individuals following vaccination with triple doses of BNT162b2. Consistent with other reports ^23-31^, these experiments showed a decrease of neutralization sensitivity of the Delta sub-lineages (both AY.36.1 and AY.122) compared to the Alpha variant (B.1.1.7) and the non-VOCs B.1.177, but higher sensitivity compared to the Omicron variant (BA.1) (Figure 7A). The two Delta sub-lineages AY.122 and AY.36.1 displayed similar neutralization sensitivity (Figure 7A).

**Figure 7:**
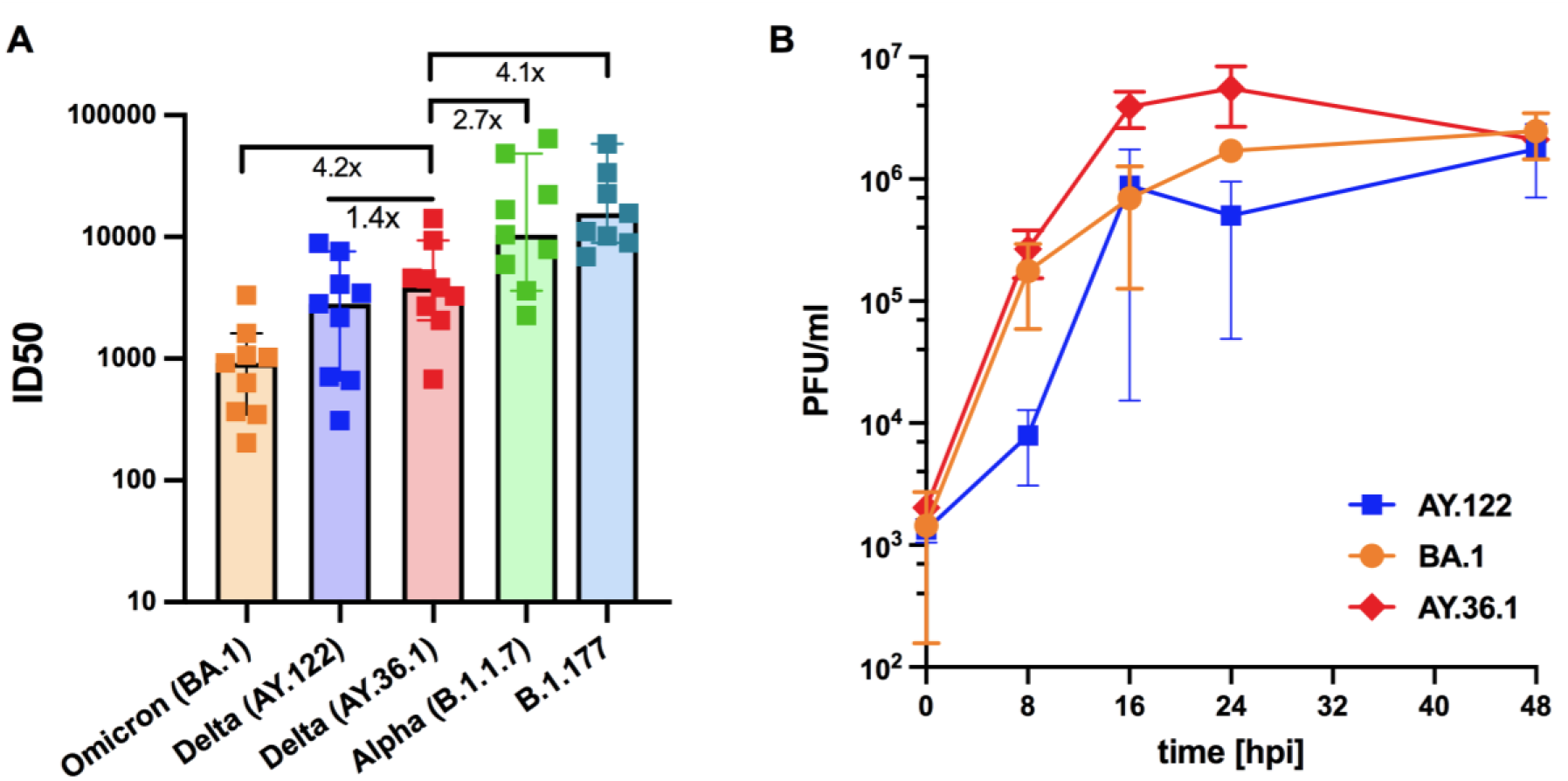
Neutralization efficacy and growth kinetics of AY.36.1 in comparison with multiple other SARS-CoV-2 variants. **(A)**. Neutralization efficacy of sera from individuals following vaccination with triple doses of BNT162b2 (*n* = 9, BNT162b2) against active virus of several VOCs variants including the Alpha (B.1.1.7), Delta (AY.122 and AY.36.1), and Omicron (BA.1) variants. B.1.177 is a non-VOC variant wildly spread in winter 2020 and early 2021. ID_50_, the serum dilution required for 50% virus inhibition. Bars represent the median ID_50_ values with 95% confidence interval. **(B)**. Growth kinetics comparing AY.36.1 with AY.122 and Omicron variant BA.1 on Calu 3 cells as titrated by plaque assay. All data represent at least two independent experiments, each with two technical replicates.

As the circulation period of AY.122 and BA.1 in Saxony overlapped with AY.36.1, we compared the replication ability of AY.36.1 with AY.122 and BA.1. For that, we infected lung epithelial cell line Calu-3 with these three variant isolates. AY.36.1 showed a clear replication advantage compared to the Delta sub-lineage AY.122, especially in the first 24 hours after infection, but no clear advantage compared to the later dominating Omicron sub-lineage BA.1 (Figure 7B). These data support higher replication rate of AY.36.1, corresponding to faster spread of it over other co-circulating Delta sub-lineages during the same period.

## Discussion

This study has revealed that long-lasting community transmission clusters developing since early autumn contributed to the incidence surge in late autumn 2021 in a cross-border region between Germany and the Czech Republic. These clusters were mostly formed by community transmission of variants imported during the travel season. In addition to imported variants, one locally occurring new variant with replication advantages played an important role as well in driving the local pandemic development possibly with an acute effect.

In this study, with available meta data we were able to investigate the transmission pattern of community transmission clusters. Analysis of two representative long-lasting AY.122 community transmission clusters in Saxony, Germany and in Ustecky Kraj, Czech Republic revealed a similar transmission pattern in the dynamic changes of geographic distribution and age distribution. In particular, a shift in age distribution that the ratio of people aged above 60 yeas increased in the later stages of community transmission was observed, which is consistent with epidemiology data that more cases of COVID-19 in elder people were often observed in the later stages of an epidemic ^32^, and also corresponding to the results of another investigation in USA that the 2020 autumn-winter wave in the United States was mainly driven by adults 20 to 49 years of age, with this age group contributing substantially to virus transmissions for the development of that autumn-winter wave ^33^. The cluster derived from the locally occurring variant AY.36.1 displayed a distinct transmission pattern: geographically much more widely spread in a short time and more elder people got infected from the very beginning. This means the consequence of the spreading of this variant could be: many elder people got infected in a short time, which may impose big pressure on the healthcare system, and this was what actually happened in Saxony in late 2021.

In line with multiple previous reports (e.g. ^23-31^), functional analyses revealed that the Delta sub-lineages (AY.122 and AY.36.1) were more resistant against antibody-mediated neutralization compared to the Alpha variant (B.1.1.7), but much more sensitive in comparison with the Omicron variant (BA.1). The two Delta sub-lineages AY.122 and AY.36.1 had similar neutralization sensitivity, but AY.36.1 showed a clear replication advantage compared to AY.122. Our findings indicate that immune evasion of AY.36.1 is similar to other Delta sub-lineages, suggesting that increased human-to-human transmissibility (e.g. due to increased replication in the upper respiratory tract or augmented infection of cells) might contribute to the expansion of AY.36.1. All the Delta sub-lineages were outcompeted by the Omicron variant in the beginning of 2022^34^. As also reported by several other studies^24,25,27,31,35,36^, the robust neutralization evasion by the Omicron variant indicates that the Omicron variant is more adept than the Delta to spread in populations that are vaccinated, which explains the takeover of AY.36.1 by the Omicron variant despite the early Omicron variant BA.1 has no clear replication advantage compared to AY.36.1.

Our investigation indicates that locally occurring new variants with a replication advantage or reduced sensitivity to antibody might cause a significant impact on the local pandemic development, which emphasizes the importance of regular genomic epidemiology analysis and mutant surveillance. To make the surveillance more real-time, it will be beneficial to reduce the turnaround time between sampling and sequence acquirement.

In this investigation, we were basically looking into this pandemic wave through one microscope. Information acquired through this method concurs with information achieved through analysis of large-scale data, but with more details. This is also a quite rare opportunity that we could perform the same analysis with data collected from two countries. Therefore, the information is least affected by local population, policy, difference in medical system and other factors, and might reflect a more general pattern for community transmission clusters derived from imported variants without non-pharmaceutical intervention.

Overall, our investigation indicates, to prevent severe burdens on healthcare systems caused by a sharp increase of COVID-19 incidence or the performance of certain mutant variants, it is important to monitor transmission clusters and to detect possible emerging new variants at the earliest possible time through regular genomic epidemiology analyses. These data suggest that mitigation approaches should be more effective if taken in the earlier stages of community transmission, such as by offering free tests to break the transmission chains. This could prevent or at least mitigate a dramatic incidence increase later. Furthermore, information provided by virus genomic epidemiology and genome surveillance, as well as other surveillance measures such as wastewater surveillance, may facilitate the accomplishment of appropriate prevention/mitigation actions and therefore help keeping normal daily life in most time.

## Data Availability

All data produced in the present work are contained in the manuscript.

## Acknowledgements

We thank all researchers who are working around the clock to generate and share genome data on GISAID (http://www.gisaid.org). We specifically thank colleagues at the Institute of Medical Microbiology and Virology, University Hospital Carl Gustav Carus, for their work in performing SARS-CoV-2 sample testing and sequencing sample preparing, and we thank the local testing labs in Saxony for their support for collecting sequencing samples. We thank the Dresden concept Genome Centre for their sequencing efforts. We thank the Robert Koch Institute for the data management and sharing. Parts of this study were supported by a grant from the German Ministry of Health (BMG) to A.D. (project LüSeMut) as well as by a grant from the State Parliament of the Free State of Saxony to A.D. B.Y. is in part supported by a funding from German Research Foundation (DFG YI175/1-1). We thank Dr. med. Robin R. Weidemann and A. Zabzinski for help with this project. We thank all the collaboration partners who contributed to the project LüSeMut.

## Data availability

A list of GISAID accession ID for AY.36.1 samples that strictly match the definition of AY.36.1 as described at https://github.com/cov-lineages/pango-designation/issues/434 containing both the three AY.36 signature mutations (S: 1104L, orf1b:721R, orf1b:1538L) and the four extra mutations of AY.36.1 (orf9b: 3S, orf3a: 223I, orf1a: 944L, N: 6L) is provide at https://github.com/genomesurveillance/delta-variant-sublineage. The processed SARS-CoV-2 AY.36.1 genome data in the form of phylogenetic tree are also available at https://github.com/genomesurveillance/delta-variant-sublineage. More general information about AY.36.1 genome number in each country during certain time period can be acquired by choosing the relevant location and collection period on the GISAID database (with searching items: VOC Delta (variants); Substitutions: N_P6L, NS3_T223I, NSP3_S126L, NSP3_P1469S; Complete; Low coverage excluded; Collection date complete). To access sequence data from GISAID, registration with https://www.gisaid.org/ is necessary, which involves agreeing to GISAID’s Database Access Agreement. Biological materials (i.e. virus variant isolation) generated as a part of this study will be made available but may require execution of a materials transfer agreement.

## Code availability

Data processing and visualization was performed using publicly available software, primarily RStudio v1.3.1093. Code for constructing phylogenetic maximum likelihood (ML) and time trees as well as phylogeographic analyses is available at https://github.com/genomesurveillance/delta-variant-sublineage, which is modified from SARS-CoV-2-specific procedures github.com/nextstrain/ncov.

